# Correlates of asymmetric venous drainage in resting state functional magnetic resonance imaging data

**DOI:** 10.1101/2025.01.06.25320046

**Authors:** Nastaran Schwarz, Benedikt Sundermann, Christian Mathys

**Author notes:** Corresponding author: PD Dr. med. Benedikt Sundermann Evangelisches Krankenhaus Oldenburg Steinweg 13-17, 26122 Oldenburg, Germany. Theses authors contributed equally (Benedikt Sundermann, Christian Mathys).

## Abstract

Functional magnetic resonance imaging (fMRI) is an indirect technique for measuring cerebral neural activity. It depends on the local cerebral vasculature and perfusion. It has thus been suggested that pathologies and normal variants of the cerebral vasculature can bias conclusions drawn from fMRI. The purpose of this study was to assess whether frequently observed asymmetries of the transverse sinuses are associated with apparent asymmetries of fMRI data timecourses. We re-analysed a publicly available resting state fMRI dataset (n = 135 healthy human subjects included in the main analysis). Voxel-mirrored homotopic connectivity (VMHC) was calculated as a measure of fMRI timecourse symmetry, reflecting the correlation of signal fluctuations in mirroring voxels in both cerebral hemispheres. Transverse sinus (TS) cross-sectional areas were measured in structural MRI data. In the main analysis, we assessed whether VMHC and TS asymmetry exhibited a negative linear association in brain areas near the transverse sinus. This was supplemented by exploratory whole brain analyses, including mapping of subthreshold effects. In the main hypothesis test, VMHC was not significantly associated with TS asymmetry in brain regions close to the transverse sinuses. In the whole brain analyses, VMHC was significantly negatively associated with TS asymmetry in more distant brain areas, and subthreshold effects resembled venous drainage patterns. The results suggest that part of the variance in resting state fMRI signal fluctuations might be explained by asymmetrical venous drainage. Thus, fMRI results might be confounded by the normal variability of the human intracranial venous system.

## 1. Introduction

Functional magnetic resonance imaging (fMRI) is a widely applied clinical and scientific method to assess brain activity. However, fMRI does not measure neural activity directly but instead relies on changes in blood volume and oxygenation as indirect indicators (Drew et al., 2020; Gauthier and Fan, 2019; Ogawa et al., 1990; Tsvetanov et al., 2021). Information about venous drainage is thus strongly represented in unprocessed fMRI raw data (Tong et al., 2011; Tong et al., 2017). It has therefore been suggested that fMRI signals obtained from draining veins may bias conclusions about neural activity (Boubela et al., 2015; Huck et al., 2023; Kalcher et al., 2015; Turner, 2002). Functional connectivity (FC) analyses based on resting-state fMRI (rs-fMRI) are considered particularly vulnerable to a wide range of biases including physiological pulsations (Birn, 2012; Chu et al., 2018) given the absence of an explicit task model. It has been further suggested that functional networks derived from rs-fMRI partially reflect vascular anatomy rather than neuronal connectivity (Tong et al., 2015) and correlations between fMRI signal timecourses in the brain and nearby large veins have been observed (Zhong et al., 2024). Accordingly, a potential bias caused by normal variants of the brain’s venous system on rs-fMRI has been reported: Developmental venous anomalies mimicked clusters of brain activity in FC-based maps of functional brain networks (Sundermann et al., 2018). Beyond such localized anomalies of venous drainage, venous drainage patterns are generally variable in the normal population. Asymmetry of the transverse sinuses (TS) is a particularly frequent and variable finding (Iwanaga, 2020). For example, a venographic magnetic resonance imaging study revealed transverse sinus asymmetries (> 10% diameter difference) in 50.6% of patients with migraine. Of these, 16.9% were classified as moderate and 24.1% as severe while 9.6 % exhibited unilateral aplasia (Fofi et al., 2012). Relative dominance of the right TS is observed more commonly (Alper et al., 2004; Ayanzen et al., 2000; Fofi et al., 2012; Goyal et al., 2016; Iwanaga, 2020; Manara et al., 2010; Rollins et al., 2005; Surendrababu et al., 2006; Widjaja and Griffiths, 2004). Potential biases caused by the TS have been discussed particularly in the field of higher visual cortex research with stimulation-based fMRI (Winawer et al., 2010).

Aim of this study was to assess whether macroscopic variability of cerebral venous drainage in terms of asymmetric TS is reflected in rs-fMRI data time courses used for FC analyses after state-of-the-art preprocessing. We used voxel-mirrored homotopic connectivity (VMHC) mapping (Zuo et al., 2010) as a surrogate for determining asymmetry in rs-fMRI data: The advantages of this technique are its relative simplicity (voxel-wise FC calculation between mirrored voxels in both hemispheres) and the independence from pre-defined seed regions of interest (ROIs). Even though VMHC as originally devised (Zuo et al., 2010) is intended to only capture homotopic connectivity (HC) omitting non-homotopic FC, it can more generally capture time course asymmetries of typically preprocessed rs-fMRI data.

The following hypothesis should be tested: Higher TS asymmetry (ratio of cross-sectional areas) is associated with decreased symmetry in FC estimates (i.e. negative linear association with VMHC). Such an association is mainly expected in posterior parts of the brain (< 4 cm distance from the TS). The hypothesis-driven main analysis was supplemented by exploratory analyses including a whole-brain analysis, the assessment of underlying spatial distributions in unthresholded VMHC maps as well as group comparisons (dichotomization based on TS asymmetry).

## 2. Materials and Methods

### 2.1. Structural and functional MRI dataset

This study is based on data from the Leipzig Study for Mind-Body-Emotion Interactions (LEMON) (Babayan et al., 2019), publicly available in anonymized form through the OpenNeuro database (Markiewicz et al., 2021) (https://doi.org/10.18112/openneuro.ds000221.v1.0.0). Only the dataset version published under a public domain (CC0) license, i.e. not including additional behavioral data, was used in this analysis. The LEMON study was carried out in accordance with the Declaration of Helsinki, and its study protocol was approved by the ethics committee at the medical faculty of the University of Leipzig (reference number 154/13-ff). Participants included in the study provided written informed consent prior to any data acquisition for the study, including consent for their data to be shared anonymously (Babayan et al., 2019). As confirmed by the local ethics committee at the medical faculty of the University of Oldenburg, formal approval was not necessary for this secondary data analysis (waiver reference number: 2021-165).

The original dataset comprised n = 227 healthy subjects in a young (20 to 35 years) and older (59 to 77 years) subgroup. Primary inclusion and exclusion criteria are described in detail in the original publication (Babayan et al., 2019). We carried out additional data quality assessment during structural and functional MRI data analysis and excluded subjects because of incomplete data, structural abnormalities, based on pre-processing outputs, including excessive head movement (see below). To avoid unnecessary heterogeneity in the data, we additionally restricted the analyses to subjects with right-dominant or co-dominant TS by excluding subjects with the less common left-dominant TS (see Supplementary Fig. 1 for detailed information on subject selection). The final dataset in the main analysis consisted of 135 subjects. Further exploratory analyses were carried out in a subgroup (n = 113) excluding participants with moderate TS asymmetry (see Supplementary Fig. 1).

Brain MRI data were acquired at 3 Tesla (Magnetom Verio, Siemens, Erlangen, Germany) with a 32-channel head coil. Analyses are based on the following unprocessed MRI data: 1) resting state fMRI (T2*-weighted gradient-echo EPI BOLD, axial acquisition orientation, phase encoding direction = A > P, in-plane pixel size = 2.3 x 2.3 mm, field of view = 202 mm, imaging matrix = 88 × 88, 64 slices with 2.3 mm thickness, repetition time = 1400 ms, echo time = 30 ms, flip angle = 69°, echo spacing = 0.67 ms, bandwidth = 1776 Hz / pixel, partial fourier 7/8, no pre-scan normalization, multiband acceleration factor = 4, 657 volumes, slice order = interleaved, overall duration = 15 min 30 s), 2) MP2RAGE (Marques et al., 2010) (sagittal acquisition orientation, one 3D volume with 176 slices, repetition time = 5000 ms, echo time = 2.92 ms, inversion time 1 = 700 ms, inversion time 2 = 2500 ms, flip angle 1 = 4°, flip angle 2 = 5°, pre-scan normalization, echo spacing = 6.9 ms, bandwidth = 240 Hz/pixel, field of view = 256 mm, voxel size = 1 mm isotropic, GRAPPA acceleration factor 3, slice order = interleaved, overall duration = 8 min 22 s) (Babayan et al., 2019).

### 2.2. Transverse sinus asymmetry quantification

The nearly triangular structure of the TS on the sagittal T1-weighted MP2RAGE image was used to calculate the Cross-Sectional Area (CSA) of the left and right TS, CSA_TS_. The CSA_TS_ 2.5 cm lateral to the confluence of sinuses (Torcular Herophili) (Granger and Tubbs, 2020) was measured using the MITK Workbench (http://www.mitk.org/, version v2021.02) as illustrated in Fig. 1.

**Fig. 1.**
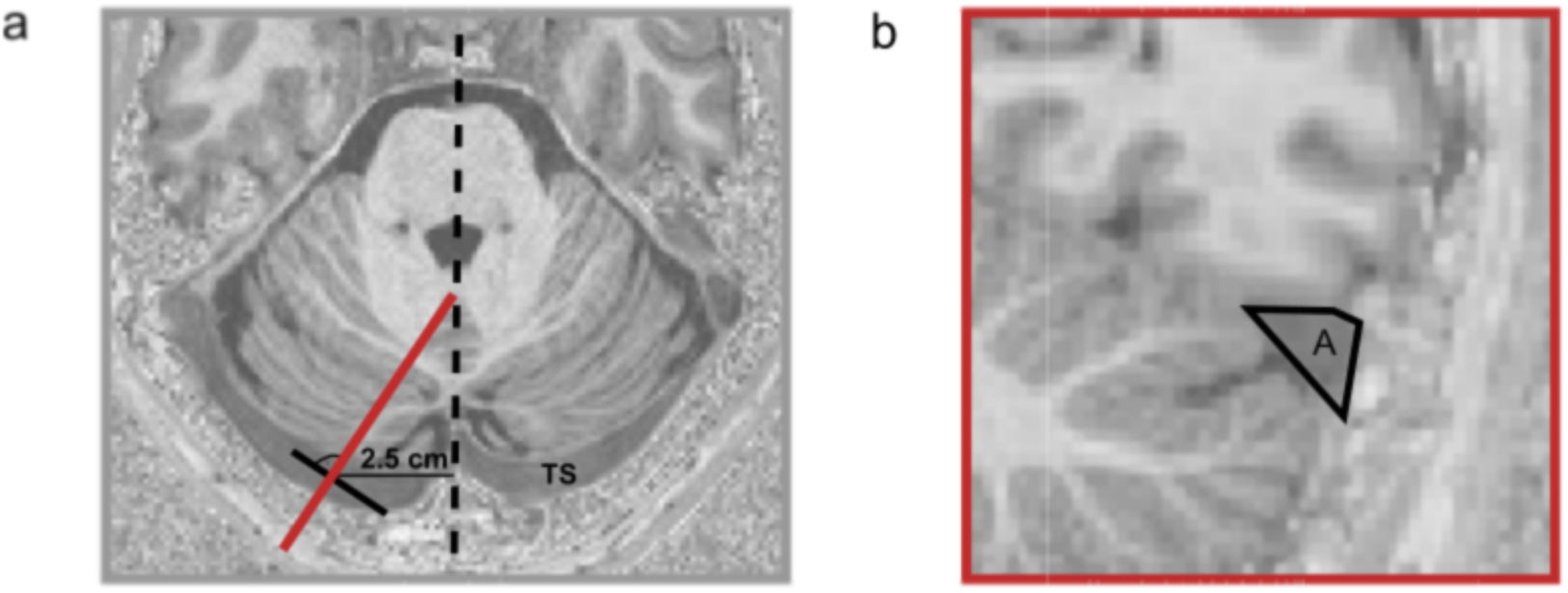
Illustration of the measurement principle and position of transverse sinus cross-sectional areas underlying the quantification of transverse sinus asymmetry. The para-sagittal measurement plane b) is defined orthogonally (red line) to the center line (black line) of the transverse sinus, 2.5 cm laterally to the midpoint of the sinus confluence in the axial plane a).

The CSA_TS_ of each side, calculated individually for each patient, was compared against each other, which resulted in the asymmetry measure (CSA_TS_ ratio). The CSA_TS-right_ was always divided by the CSA_TS-left_. This avoided heterogeneity in the data. Subjects with a left dominant type and a CSA_TS_ ratio < 1 were excluded from subsequent analyses, thus limiting patients with TS asymmetry to the more common right-dominant form.

### 2.3. fMRI preprocessing and VMHC calculation

An individual 3D T1-weighted image with properties comparable to MPRAGE (i.e., dark background) was required as an anatomical reference for fMRI preprocessing. It was calculated for each subject by multiplying the MPRAGE uniform ‘T1w’ and second inversion image ‘inv2’ with the FMRIB software Library (FSL) utilities (Jenkinson et al., 2012). Main MRI data preprocessing was carried out using fMRIPrep (Esteban et al., 2019) (version 20.0.7), briefly consisting of motion estimation and correction, co-registration of fMRI and structural MRI data, estimation of noise regressors, as well as standard space normalization. Detailed information is presented in the Supplementary Methods.

Subsequent denoising was carried out using fMRIDenoise (Finc, 2021) (version 0.2.1) analogous to two of our previous studies (Sundermann et al., 2023; Sundermann et al., 2024a). This comprised regressing out 24 head motion parameters (3 translations, 3 rotations, their 6 temporal derivatives, and their 12 quadratic terms) (Satterthwaite et al., 2013), 8 physiological noise parameters (mean physiological signals from white matter and cerebrospinal fluid, their 2 temporal derivatives, and 4 quadratic terms) (Satterthwaite et al., 2013), as well as movement spike regression based on frame-wise displacement (FD > 0.5 mm) and so-called “DVARS” (> 3) thresholds (Power et al., 2012), temporally filtering (0.008 – 0.08 Hz), and smoothing the resulting standard space image with a Gaussian kernel (FWHM = 6 mm). This preprocessing leads to denoised fMRI data in a common standard space as input for further analyses.

VMHC (Zuo et al., 2010) maps were obtained by calculating maps with z-transformed Pearson correlation coefficients of timecourses of pairs of mirrored voxels in both hemispheres using the Data Processing Assistant for Resting-State fMRI (DPARSF, version 5.2) (Chao-Gan and Yu-Feng, 2010), based on Matlab 2019b (The MathWorks, Natick, MA, USA).

### 2.4. Analyses of an association between VMHC and transverse sinus asymmetry: general aspects

Group analyses of VMHC maps (dependent variable) were carried out using general linear models in SPM12 (https://www.fil.ion.ucl.ac.uk/spm/). Given that VMHC maps are symmetric by definition and thus redundant information is present in both hemispheres, analyses were restricted to voxels with at least 10% gray matter (GM) probability in a single hemisphere by masking. The mask was created based on the GM mask corresponding to the standard space used during preprocessing, obtained from TemplateFlow (Ciric et al., 2022).

### 2.5. Analyses of an association between VMHC and transverse sinus asymmetry: hypothesis-driven main analysis

An SPM second-level analysis with a multiple linear regression design was carried out for the primary hypothesis in the main analysis sample. The z-transformed VMHC maps were used as dependent variables. The CSA_TS_ ratios (CSA_TS-right_ / CSA_TS-left_) were the independent variables. Age and sex were included as covariates and a constant term was included in the model. The hypothesis-driven main analysis was restricted to a single hemisphere (see general description above) and it was limited to gray matter regions located a maximum of 40 mm from the transverse sinus, which was manually delineated in the MNI template (see Fig. 2). A t-contrast was defined to reflect a negative association of the CSA_TS_ ratios with VMHC. The hypothesis-driven analysis was corrected for multiple comparisons using the family-wise error rate (FWE) at the alpha = 0.05 level at the voxel-level.

**Fig. 2.**
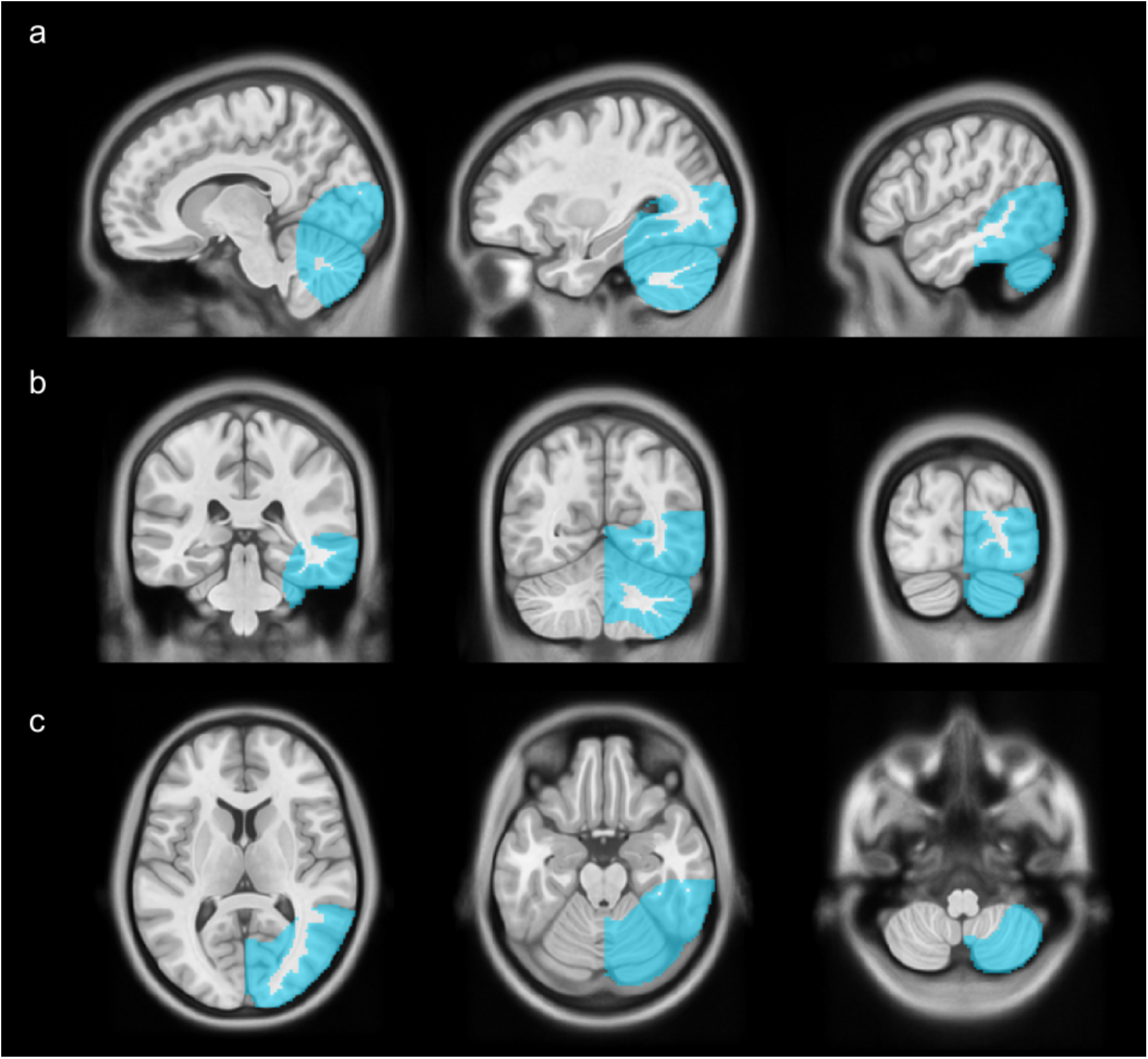
Illustration of the grey matter mask (blue) around the transverse sinus (maximum distance: 4 cm, one hemisphere only) used in the hypothesis-driven main group analysis of the symmetrical VMHC maps in a) sagittal, b) coronal, and c) axial orientation overlaid on the MNI standard space template.

### 2.6. Analyses of an association between VMHC and transverse sinus asymmetry: additional exploratory analyses

We carried out a linear regression analysis identical to the main hypothesis-driven one but not restricted to the region around the transverse sinuses. Considering the symmetry of the VMHC maps, the analysis was restricted to a single-hemisphere gray matter mask (as described above).

In addition to thresholding (test of significance), we represented and visually interpreted the underlying unthresholded contrast maps. The rationale for interpreting unthresholded maps is that they might show more interpretable global venous patterns associated with significant (i.e., above-threshold) peaks. They might also suggest entirely subthreshold venous patterns as a starting point for future research.

As part of a secondary exploratory analysis, subjects were divided into a group with mostly symmetrical CSA_TS_ ratio between the left and right side < 1,3) vs. substantially asymmetrical drainage of the more common right-dominant type CSA_TS_ ratio between the sides > 1.6). Subjects with intermediate CSA_TS_ asymmetry (i.e., CSA_TS_ between 1.3 and 1.6 % differences) were not included in the analyses (see the “additional group-based analysis sample” in supplementary figure 1). Rationale for this group-based analysis is to assess the robustness of main findings (two secondary analysis models for similar research questions) and to reduce potential outlier influences (i.e. of cases with unilateral near-aplasia). For this purpose, an SPM second-level analysis was carried out with a 2-sample t-test design, with which the two groups were compared. Age and sex were again included as covariates. The analysis was again limited to a single-hemisphere gray matter mask (as described above). A t-contrast for no relevant asymmetry was defined.

## 3. Results

### 3.1. Sample characteristics and TS asymmetry

The main analysis sample (n = 135) comprised 48 (35.3%) female and 87 (64.7%) male participants with an average age of 36.1 years (with a bimodal distribution due to the original LEMON study inclusion criteria (Babayan et al., 2019)). The CSA_TS_ ratio was on average 2.16 (standard deviation: 1.61, range: 1 to 11.91) in line with a right-sided TS predominance.

The additional group-based analysis sample (n = 113) comprised 40 (35.4%) female and 73 (64.6%) male participants with an average age of 35.6 years. 77 participants (68.1%) exhibited high and 36 participants (31.9%) exhibited no or low TS asymmetry.

### 3.2. Analyses of an association between VMHC and transverse sinus asymmetry: hypothesis-driven main analysis

In the main linear regression analysis of VMHC and TS asymmetry, restricted to brain regions within 4 cm distance of the transverse sinus and conservative FWE-control for multiple comparisons, no statistically significant effect was observed.

### 3.3. Analyses of an association between VMHC and transverse sinus asymmetry: additional exploratory analyses

In the linear regression analysis, not restricted to areas around the transverse sinuses but otherwise identical to the main analysis, we observed a significant negative association of TS asymmetry and VMHC (i.e., higher TS symmetry was associated with higher VMHC, representing higher symmetry of BOLD-timecourses) in the area of the Sylvian fissure (Fig. 3a). Visualization of the underlying unthresholded contrast map pointed towards potential further subthreshold effects (Fig. 3b) not surviving FWE-based correction for multiple comparisons.

**Fig. 3.**
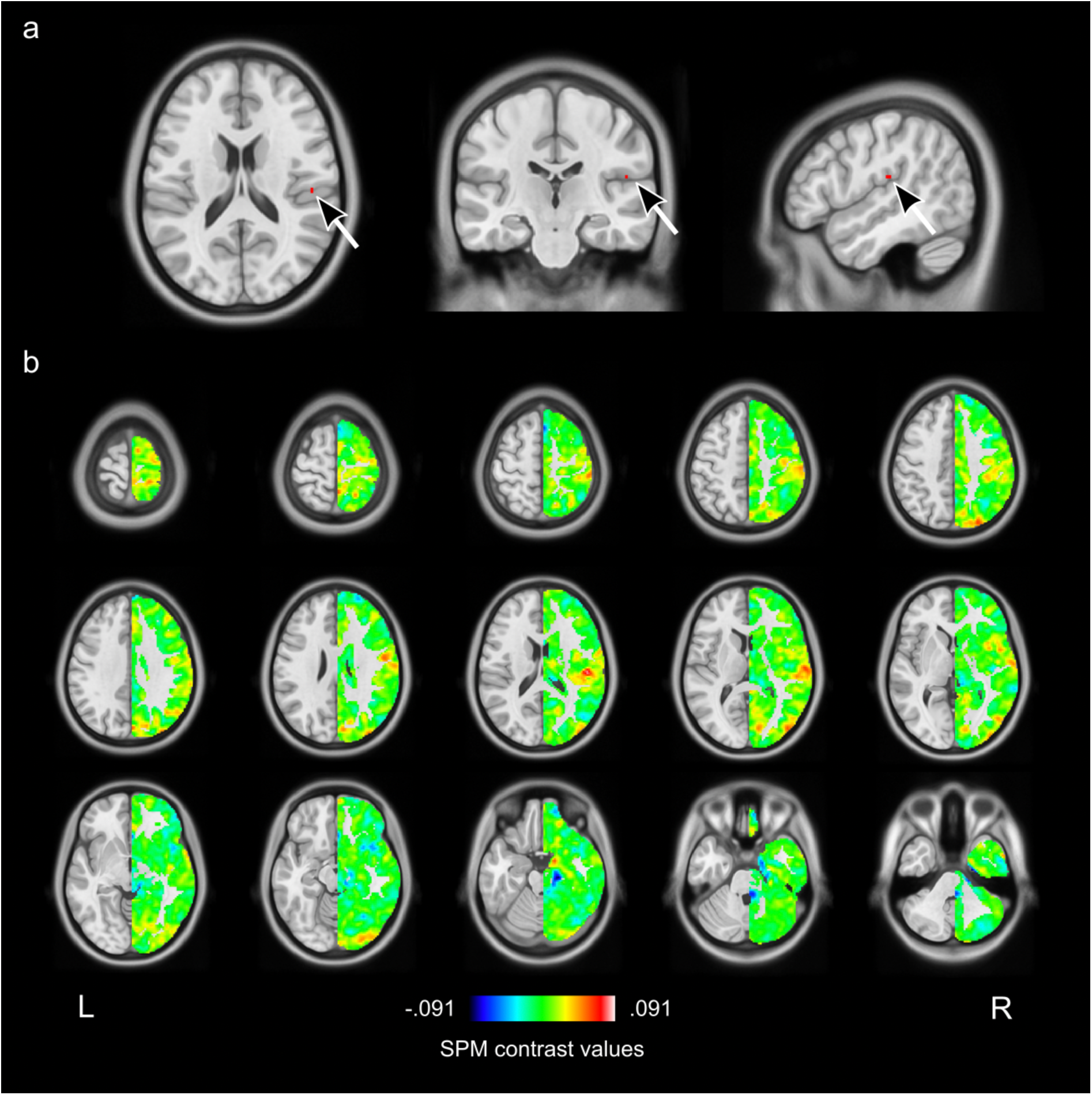
Whole brain voxel-wise association (linear regression) of transverse sinus asymmetry and VMHC, contrast: negative effect of TS asymmetry: a) thresholded whole brain results showing a significant association in the Sylvian fissure (p < 0.05, FWE-corrected, cluster size: 2 voxels, MNI peak coordinates: x = 50, y = −22, z = 18), b) unthresholded results (SPM contrast maps)

In the group-based analysis (dichotomized: no/low TS asymmetry vs. strong TS asymmetry), we observed no significant group effect on VMHC with FEW-correction for multiple comparisons. Side-by-side visualizations of unthresholded contrast maps underlying the regression-based main analysis and the group-based additional analysis are presented in figure 4. Both show effects resembling venous structures (see, for example, the location of the basal vein of Rosenthal) as well as antero-posterior patterns (stronger in the group-based analysis).

**Fig. 4.**
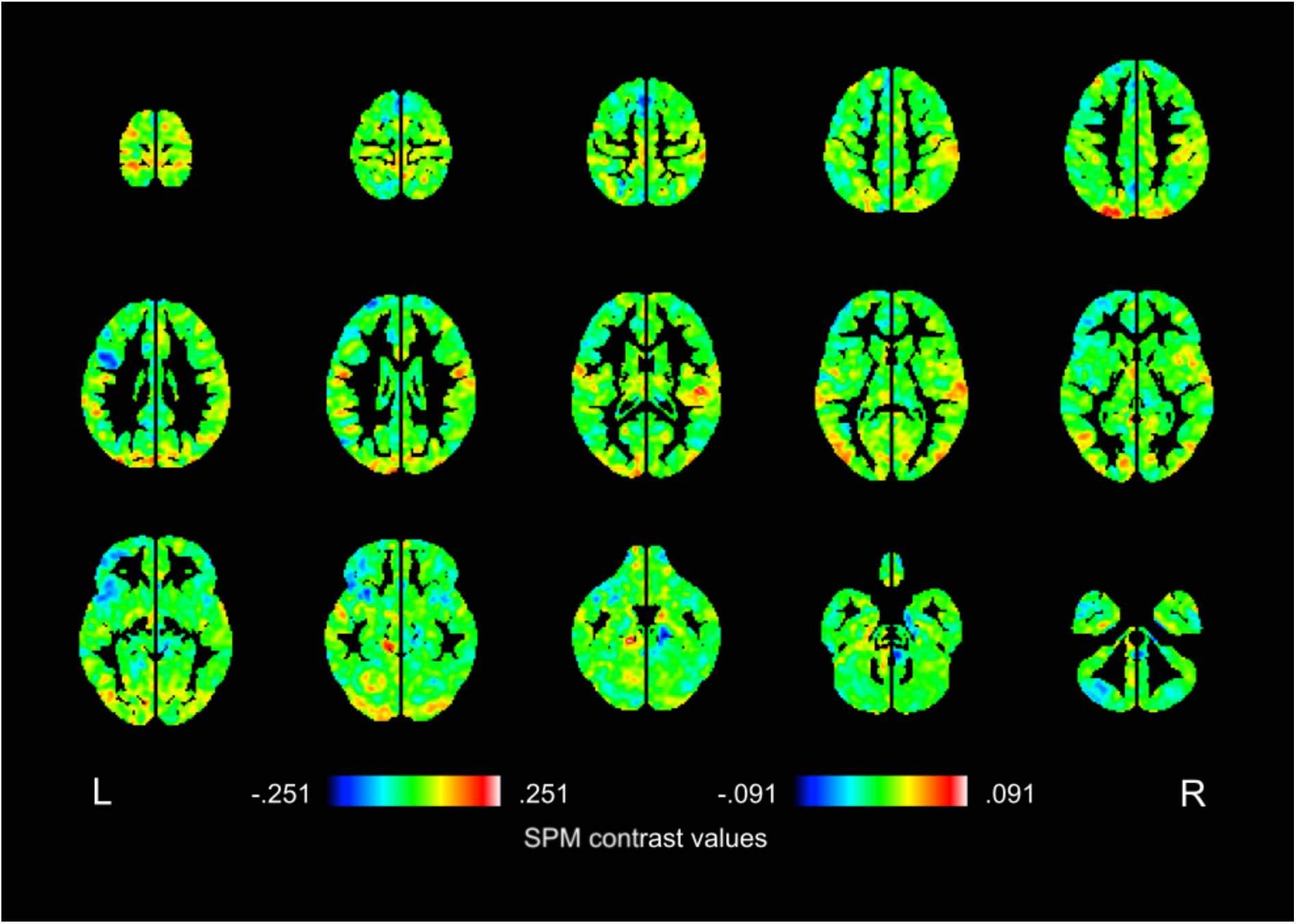
Whole brain voxel-wise association of transverse sinus asymmetry and VMHC. Comparison of unthresholded results (SPM contrast maps) between the main analysis (linear regression, contrast: negative effect of TS asymmetry), shown on the right (R), and the exploratory group comparison (contrast: positive effect of no/low vs. high TS asymmetry), shown on the left (L). Results from both analyses mainly exhibit an anterior-posterior pattern (with few exceptions in large veins such as the basal veins of Rosenthal), largely depending on the distance from the transverse sinuses, which is more obvious in the group-based analysis.

## 4. Discussion

To summarize, brain areas in close proximity (< 4 cm distance) to the TS did not exhibit statistically significant associations between the asymmetry of rs-fMRI signal fluctuations measured by VMHC and TS asymmetry (main hypothesis test with conservative multiple comparison correction). However, exploratory analyses including more distant grey matter regions and subthreshold effects point towards an association of TS asymmetry and fMRI data asymmetry. Findings are thus generally in line with previous research suggesting an association of intracranial venous anatomy with findings in task-based and resting-state fMRI (Boubela et al., 2015; Kalcher et al., 2015; Sundermann et al., 2018; Zhong et al., 2024).

In the linear regression analysis extending the main hypothesis-driven analysis to the entire grey matter, a statistically significant cluster was observed in the Sylvian fissure (Fig. 3a) even despite conservative multiple comparison correction. This observation is noteworthy, since it highlights that associations with venous architecture in typical statistical testing regimes for fMRI (Han and Glenn, 2018; Monti, 2011) can appear “blob-like” and can thus resemble findings typically interpreted as local connectivity effects (or possibly local activation) in fMRI research (Cremers et al., 2017). The localized finding in this analysis, despite distant from the TS, is anatomically meaningful, since Sylvian fissure veins typically drain into the TS (entry usually lateral to the point of measurement in this study) via Labbé’s Inferior Anastomotic Vein (LV) either directly or via tentorial sinuses (Minca et al., 2022; Tomasi et al., 2021). The LV has a variable anatomical course (Minca et al., 2022). Considering potential anatomical variability, the observed association of TS asymmetry and VMHC in the Sylvian fissure in this study might thus be attributed to the fact that if the TS is smaller on one side, the blood from the Sylvian fissure would have to be distributed via different drainage routes. Research on anatomical variability of the drainage pathways including the LV has been focused on patterns and landmarks related to neurosurgical approaches (e.g. (Bigelow et al., 1993; Minca et al., 2022; Tomasi et al., 2021)). We could not identify a previous study quantitatively assessing associations of LV and medial TS dimensions.

Looking beyond what appears to be localized effects, assessment of unthresholded result maps (Sundermann et al., 2024b; Taylor et al., 2023) reveals an anterior-posterior gradient (i.e. varying with proximity to the TS) with a tendency towards stronger negative relationship between TS asymmetry and VMHC in occipital areas (Fig. 3b and 4). Larger venous structures such as the basal vein of Rosenthal, which has been discussed as a potential confounder of fMRI activation attributed to the amygdala (Boubela et al., 2015), can be visually recognized both in the linear regression analysis and in the group-based analysis (Fig. 3b and 4), albeit partially exhibiting opposite effects in both analyses. In addition to effects assumed to be more directly associated with TS asymmetry alone, the observed global patterns might be influenced by the non-uniform global intracranial distribution of vascular pulsations present in BOLD fMRI data (Raitamaa et al., 2021) or more general venous vessel probability (for detailed mapping see (Bernier et al., 2018)). It has also been recently demonstrated that homotopic FC estimates can be influenced by local vascular and systemic arterial factors (Schneider et al., 2023). Furthermore, associations of the BOLD signal with venous asymmetry might be influenced by still not completely understood effects of neurovascular coupling, including potential effects of pressure gradients (Drew, 2022).

In addition to representing direct vascular effects, it is also conceivable that brain structural asymmetry (e.g. gyral patterns) is associated with asymmetries of the cerebral venous vasculature ranging from minor local displacement to more regional effects of joint development. For example, regional vascular density is positively correlated with cortical thickness (Bernier et al., 2018) and an association of cortical microstructure and venous architecture has recently been reported as a preprint (Knoll et al., 2023). Joint development of intracranial vessels and other anatomical structures with presumed mutual influences has long been established in embryology (Padget; Raybaud, 2010) and recent functional neuroimaging research suggests close organizational relationships between cerebral vascular system functioning and resting-state networks (Bright et al., 2020). Thus, differences in apparent functional homotopy (measured by VMHC) might in part reflect minor structural brain symmetry variations.

Findings of this study raise caution, that the variability of macrovascular cerebral venous drainage patterns might bias results obtained from fMRI using standard preprocessing and statistical analysis approaches. VMHC is used as a relatively simple surrogate marker of general rs-fMRI timecourse asymmetry in such a common late intermediate analysis stage to facilitate relatively wide generalizability across rs-fMRI studies. While venous effects on BOLD fMRI raw data are generally well-known and inherent to its measurement principle (Drew et al., 2020; Gauthier and Fan, 2019; Ogawa et al., 1990; Tsvetanov et al., 2021), this has predominantly been discussed in terms of pulsations and corresponding fMRI denoising strategies, limited by temporal resolution of the fMRI data and potential aliasing (Birn et al., 2014; Glover et al., 2000; Strother, 2006). The high inter-subject variability and asymmetry of large veins and venous sinuses, however, has received little attention in the fMRI research community. Visual classification of drainage patterns, asymmetry quantification, or detailed venous mapping is not typically included in fMRI analyses for example in psychology, cognitive and clinical neuroscience. Conventional denoising strategies to some extent addressing physiological pulsations more generally (Birn et al., 2014; Esteban et al., 2019; Glover et al., 2000; Strother, 2006) might thus be supplemented by data on anatomical venous drainage variability including normal variants. This might be of particular interest in relatively small samples prone to outlier effects (Cremers et al., 2017; Wager et al., 2005; Woolrich, 2008). Denoising techniques based on independent component analyses (ICA) such as FSL FIX (Salimi-Khorshidi et al., 2014) or principal component analysis (PCA) such as CompCor (Behzadi et al., 2007) might be of particular interest since some degree of venous drainage variability might already be included in predominantly venous components to be removed from the fMRI raw data during preprocessing. Therefore, evaluating the particular benefit of ICA- or PCA-based denoising is a potential line of future research in this area. Recently, a model for correcting venous biases based on high-resolution venous system mapping has been proprosed for 7 Tesla whole brain fMRI (Huck et al., 2023). Further dedicated venous modeling and artifact reduction strategies are being developed in ultra-high resolution fMRI (e.g. (Kay et al., 2020)), which might in the future be translated to more routine human whole brain fMRI acquisitions in e.g. clinical and cognitive neuroscience.

### 4.1. Potential limitations

In this retrospective analysis of a single functional imaging modality (rs-fMRI), it cannot be distinguished if the associations of VMHC and TS asymmetry represent purely vascular effects in the BOLD fMRI data without a neuronal basis, true functional activity differences (i.e. representing altered brain organization associated with variability of venous drainage pathways), or a combination of these potential effects. Future multi-modal studies for example using combined EEG-fMRI (Diukova et al., 2012; Mele et al., 2019; Tsvetanov et al., 2021), calibrated fMRI (Chen and Gauthier, 2021; Hoge, 2012; Tsvetanov et al., 2021) or fMRI techniques potentially less affected by macrovascular venous signal contributions such as VAscular-Space-Occupancy (VASO) (Akbari et al., 2023), or BOLD-fMRI with enhanced arterial contrast (Priovoulos et al., 2023) or phase-based venous suppression (Curtis et al., 2014) may help resolve this issue.

Brain structure including gyral patterns (Chen et al., 2022; Chiarello et al., 2016; Kong et al., 2018; Roe et al., 2022) and subcortical structures (Gomez-Ramirez and Gonzalez-Rosa, 2022) is slightly asymmetric reflected by an asymmetry of standard space templates typically used in fMRI group studies (Watkins et al., 2001). An example is the slight rotational deformation of the base of the brain (Thiebaut de Schotten and Beckmann, 2022), which can also be observed in the MNI template. The original VMHC implementation includes registration to an artificially symmetrized template (Zuo et al., 2010). We did intentionally not include registration to a symmetrized template, in order not to discard underlying asymmetries in the raw data. This limits the comparability of these results with VMHC studies aiming at FC between exactly homotopic cortical areas. In this study, we rather used the VMHC measure as a more general surrogate marker of asymmetric signal fluctuations in the preprocessed fMRI data. We thus believe that results are more generalizable to the larger number of fMRI studies using slightly asymmetric standard space templates. Despite this general approach, observed effects in preprocessed fMRI data might not be generalizable to all conceivable fMRI analyses, e.g. those using different preprocessing approaches, such as denoising based on component analyses (Behzadi et al., 2007; Pruim et al., 2015; Salimi-Khorshidi et al., 2014), or for example intra-hemispheric FC analyses (Krupnik et al., 2021). For a related discussion on asymmetrical vs. symmetrical parcellations/atlases in rs-fMRI see also (Yan et al., 2023).

The main linear regression analysis can potentially be affected by outliers, i.e. subjects with an almost absent hypoplastic TS. Despite an anatomically meaningful association, we are thus cautious regarding generalizability of the exact location of the observed local peak. However, both the correlation and group-based analysis (potentially less affected by outliers) point towards meaningful subthreshold effects (see (Sundermann et al., 2024b) for a more general discussion of the potential relevance of fMRI subthreshold effects) of venous asymmetries on rs-fMRI data.

## 5. Conclusions

While the main hypothesis test restricted to gray matter regions in close proximity to the transverse sinuses resulted in a negative result, further exploratory analyses point towards an association of higher venous drainage asymmetry with predominantly higher asymmetry of the fMRI data time courses and resembling anatomically meaningful patterns. This suggests, that part of the variance of typically preprocessed resting state fluctuations might be explained by asymmetrical venous drainage. These findings raise caution that conclusions about asymmetry of brain activity measured by fMRI (and fMRI results more generally) may be confounded by normal population-variability of the human intracranial venous architecture. Venous system mapping or improving data acquisition and preprocessing techniques are thus potential avenues of research for improving the validity of fMRI research while multi-modal studies may better help understand the interactions of vascular and neural systems architecture.

## Data Availability

This study is based on data from the Leipzig Study for Mind-Body-Emotion Interactions (LEMON) (Babayan et al., 2019, Sci Data 6, 180308, DOI: 10.1038/sdata.2018.308), publicly available in anonymized form at https://doi.org/10.18112/openneuro.ds000221.v1.0.0.

## Data availability

The previously published open dataset (Babayan et al., 2019) underlying this analysis is publicly available from https://doi.org/10.18112/openneuro.ds000221.v1.0.0.

## CRediT authorship contribution statement

**Nastaran Schwarz:** Data curation, Formal analysis, Writing – original draft. **Benedikt Sundermann:** Conceptualization, Data curation, Methodology, Supervision, Writing – original draft. **Christian Mathys:** Conceptualization, Methodology, Supervision, Resources, Writing – review and editing.

## Acknowledgements

We thank those involved in acquiring and publicly sharing the underlying LEMON (Babayan et al., 2019) dataset, particularly the study participants, researchers as well as the founders and maintainers of the OpenNeuro database (Markiewicz et al., 2021). Preliminary results of this analysis have been submitted as an academic thesis as part of the longitudinal research curriculum at the Medical Faculty of the University of Oldenburg / European Medical School Oldenburg-Groningen and presented at the 2023 Annual Meeting of the European Society of Neuroradiology (ESNR) (Schwarz et al., 2023).

## Supplementary material

### Supplementary Figure

**Supplementary Figure 1:**
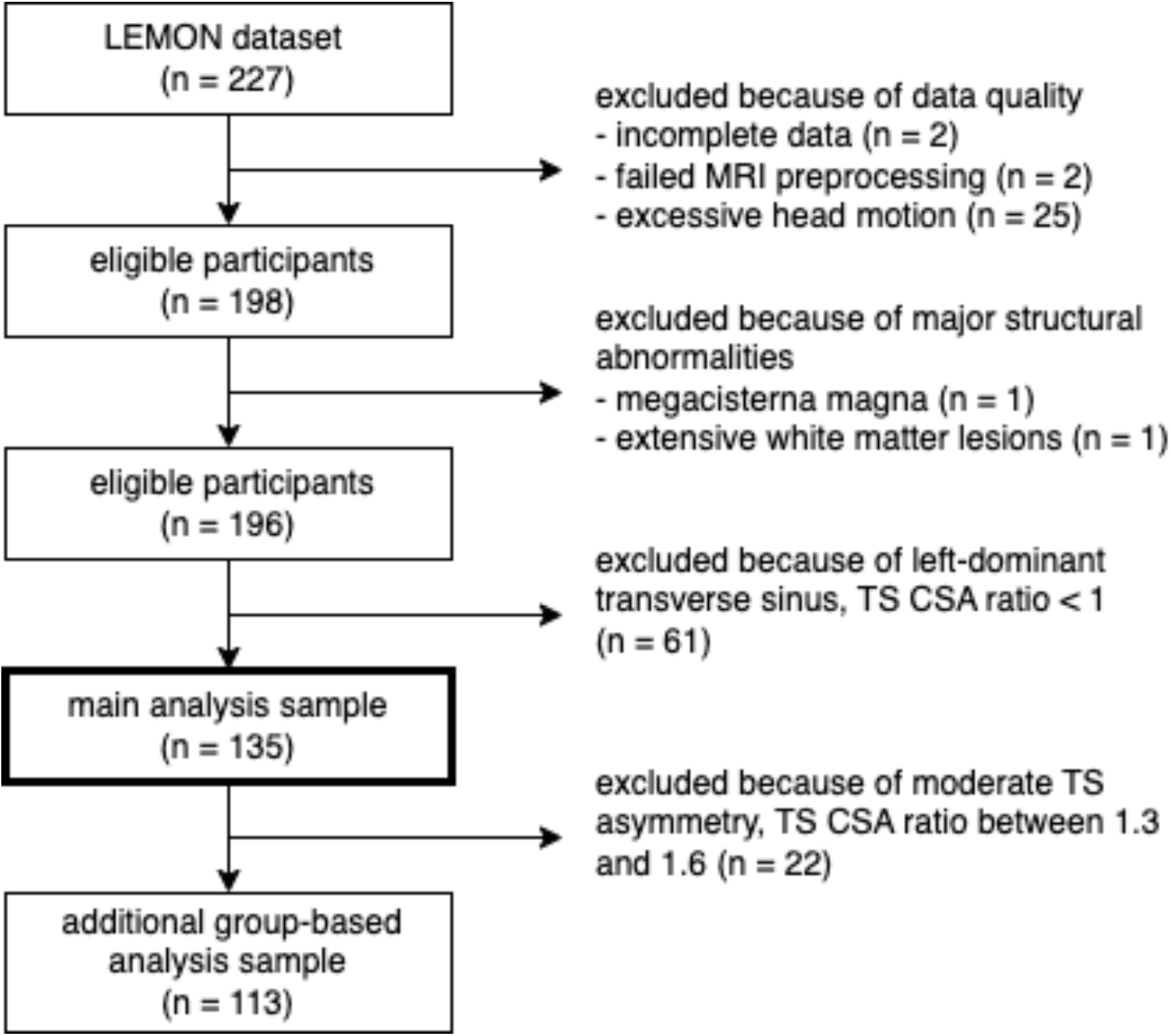
Flow chart of the included/excluded participants. Excessive head motion was defined as mean frame-wise displacement (FD) > 0,3 mm, max FD > 5mm, or >20 % outlier datapoints. TS: transverse sinus, CSA: cross-sectional area (of the left and right TS), MRI: magnetic resonance imaging.

### Supplementary Methods

#### MRI data preprocessing

The following fMRIPrep “boilerplate” describes the preprocessing of the MRI data in detail. The text generated as part of the fmriprep reports during preprocessing has intentionally been left unchanged according to the fMRIPrep recommendations (see also https://fmriprep.org/en/20.0.7/citing.html) for optimal reproducibility. As represented in this text, fMRIPrep generated multiple preprocessing outputs which could be used in different denoising and analysis strategies. Not all of these parallel outputs were used for further processing and analyses in this work. Details about which outputs were used for further modelling are presented in the main text.

“Results included in this manuscript come from preprocessing performed using *fMRIPrep* 20.0.7 (Esteban, Markiewicz, et al. (2018); Esteban, Blair, et al. (2018); RRID:SCR_016216), which is based on *Nipype* 1.4.2 (Gorgolewski et al. (2011); Gorgolewski et al. (2018); RRID:SCR_002502).

##### Anatomical data preprocessing

The T1-weighted (T1w) image was corrected for intensity non-uniformity (INU) with N4BiasFieldCorrection (Tustison et al. 2010), distributed with ANTs 2.2.0 (Avants et al. 2008, RRID:SCR_004757), and used as T1w-reference throughout the workflow. The T1w-reference was then skull-stripped with a *Nipype* implementation of the antsBrainExtraction.sh workflow (from ANTs), using OASIS30ANTs as target template. Brain tissue segmentation of cerebrospinal fluid (CSF), white-matter (WM) and gray-matter (GM) was performed on the brain-extracted T1w using fast (FSL 5.0.9, RRID:SCR_002823, Zhang, Brady, and Smith 2001). Brain surfaces were reconstructed using recon-all (FreeSurfer 6.0.1, RRID:SCR_001847, Dale, Fischl, and Sereno 1999), and the brain mask estimated previously was refined with a custom variation of the method to reconcile ANTs-derived and FreeSurfer-derived segmentations of the cortical gray-matter of Mindboggle (RRID:SCR_002438, Klein et al. 2017). Volume-based spatial normalization to two standard spaces (MNI152NLin2009cAsym, MNI152NLin2009cSym) was performed through nonlinear registration with antsRegistration (ANTs 2.2.0), using brain-extracted versions of both T1w reference and the T1w template. The following templates were selected for spatial normalization: *ICBM 152 Nonlinear Asymmetrical template version 2009c* [Fonov et al. (2009), RRID:SCR_008796; TemplateFlow ID: MNI152NLin2009cAsym], *ICBM 152 Nonlinear Symmetrical template version 2009c* [(**???**), RRID:SCR_008796; TemplateFlow ID: MNI152NLin2009cSym],

##### Functional data preprocessing

For each of the 1 BOLD runs found per subject (across all tasks and sessions), the following preprocessing was performed. First, a reference volume and its skull-stripped version were generated using a custom methodology of *fMRIPrep*. Susceptibility distortion correction (SDC) was omitted. The BOLD reference was then co-registered to the T1w reference using bbregister (FreeSurfer) which implements boundary-based registration (Greve and Fischl 2009). Co-registration was configured with six degrees of freedom. Head-motion parameters with respect to the BOLD reference (transformation matrices, and six corresponding rotation and translation parameters) are estimated before any spatiotemporal filtering using mcflirt (FSL 5.0.9, Jenkinson et al. 2002). The BOLD time-series (including slice-timing correction when applied) were resampled onto their original, native space by applying the transforms to correct for head-motion. These resampled BOLD time-series will be referred to as *preprocessed BOLD in original space*, or just *preprocessed BOLD*. The BOLD time-series were resampled into several standard spaces, correspondingly generating the following *spatially-normalized, preprocessed BOLD runs*: MNI152NLin2009cAsym, MNI152NLin2009cSym. First, a reference volume and its skull-stripped version were generated using a custom methodology of *fMRIPrep*. Several confounding time-series were calculated based on the *preprocessed BOLD*: framewise displacement (FD), DVARS and three region-wise global signals. FD and DVARS are calculated for each functional run, both using their implementations in *Nipype* (following the definitions by Power et al. 2014). The three global signals are extracted within the CSF, the WM, and the whole-brain masks. Additionally, a set of physiological regressors were extracted to allow for component-based noise correction (*CompCor*, Behzadi et al. 2007). Principal components are estimated after high-pass filtering the *preprocessed BOLD* time-series (using a discrete cosine filter with 128s cut-off) for the two *CompCor* variants: temporal (tCompCor) and anatomical (aCompCor). tCompCor components are then calculated from the top 5% variable voxels within a mask covering the subcortical regions. This subcortical mask is obtained by heavily eroding the brain mask, which ensures it does not include cortical GM regions. For aCompCor, components are calculated within the intersection of the aforementioned mask and the union of CSF and WM masks calculated in T1w space, after their projection to the native space of each functional run (using the inverse BOLD-to-T1w transformation). Components are also calculated separately within the WM and CSF masks. For each CompCor decomposition, the *k* components with the largest singular values are retained, such that the retained components’ time series are sufficient to explain 50 percent of variance across the nuisance mask (CSF, WM, combined, or temporal). The remaining components are dropped from consideration. The head-motion estimates calculated in the correction step were also placed within the corresponding confounds file. The confound time series derived from head motion estimates and global signals were expanded with the inclusion of temporal derivatives and quadratic terms for each (Satterthwaite et al. 2013). Frames that exceeded a threshold of 0.5 mm FD or 1.5 standardised DVARS were annotated as motion outliers. All resamplings can be performed with *a single interpolation step* by composing all the pertinent transformations (i.e. head-motion transform matrices, susceptibility distortion correction when available, and co-registrations to anatomical and output spaces). Gridded (volumetric) resamplings were performed using antsApplyTransforms (ANTs), configured with Lanczos interpolation to minimize the smoothing effects of other kernels (Lanczos 1964). Non-gridded (surface) resamplings were performed using mri_vol2surf (FreeSurfer).

Many internal operations of *fMRIPrep* use *Nilearn* 0.6.2 (Abraham et al. 2014, RRID:SCR_001362), mostly within the functional processing workflow. For more details of the pipeline, see <u>the section corresponding to workflows in</u> *fMRIPrep*<u>’s documentation</u>.

##### Copyright Waiver

The above boilerplate text was automatically generated by fMRIPrep with the express intention that users should copy and paste this text into their manuscripts *unchanged*. It is released under the <u>CC0</u> license.

